# Feasibility of an online antigen self-testing strategy for SARS-CoV-2 addressed to health care and education professionals in Catalonia (Spain). The TESTA’T-COVID Project

**DOI:** 10.1101/2022.04.26.22274256

**Authors:** Cristina Agustí, Héctor Martínez-Riveros, Victoria González, Gema Fernández-Rivas, Yesika Díaz, Marcos Montoro-Fernandez, Sergio Moreno-Fornés, Pol Romano-deGea, Esteve Muntada, Beatriz Calvo, Jordi Casabona

**Affiliations:** Centre of epidemiological studies on sexually transmitted infections and AIDS of Catalunya (CEEISCAT). Department of Health. Generalitat of Catalunya. Badalona. Spain; Spanish Consortium for Research on Epidemiology and Public Health (CIBERESP), Instituto de Salud Carlos III, Madrid, Spain; Germans Trias i Pujol Research Institute (IGTP), Campus Can Ruti, Badalona, Spain; PhD in Methodology of Biomedical Research and Public Health. Department of Paediatrics, Obstetrics and Gynecology and Preventive Medicine, Univ Autonoma de Barcelona, Badalona, Spain; Microbiology Department, Clinical Laboratory North Metropolitan Area, Germans Trias i Pujol University Hospital. Departament of Genetics and Microbiology, Autonomous University of Barcelona, Badalona, Spain; Occupational Health and Safety Department, Institut Català d’Oncologia (ICO), L’Hospitalet de Llobregat, Spain; Department of Paediatrics, Obstetrics and Gynecology and Preventive Medicine, Universitat Autònoma de Barcelona, Badalona, Spain

**Keywords:** SARS-CoV-2, COVID-19, Self-testing, health care professionals, education professionals

## Abstract

We aimed to implement a pilot intervention based on offering online COVID-19 selftest kits addressed to healthcare and education professionals in Spain during the peak of the 6^th^ wave caused by Omicron variant. Kits were ordered online and sent by mail, participants answered an online acceptability/usability survey and uploaded the picture of results. 492 participants ordered a test, 304 uploaded the picture (61.8%). Eighteen positive cases were detected (5.9%). 92.2% were satisfied/very satisfied with the intervention; and 92.5% found performing the test easy/very easy. We demonstrated that implementing online COVID-19 self-testing in schools and healthcare settings in Spain is feasible.

**Key findings:**

- We implemented a pilot intervention based on offering online COVID-19 selftest kits in Spain.
- We demonstrated the feasibility of the intervention during the peak of the 6th wave caused by Omicron variant.
- The intervention counted with high acceptability among healthcare and education professionals in Spain.
- Our results may contribute to define screening strategies addressed to key populations, particularly during peaks of high community transmission of SARS-CoV-2.

## Introduction

It has been estimated that nearly half of the transmissions of SARS-CoV-2 occur from asymptomatic individuals [1]. As for other infections, the screening of asymptomatic individuals at risk of being exposed to SARS-CoV-2 in order to detect and isolate infected persons early is one of the basic non-pharmaceutical preventive interventions shown to decrease incidence at the community level [2]. Antigen-detecting rapid diagnostic tests (Ag-RDTs) have been proposed as suitable tools for point-of-care screening of individuals potentially exposed. The main advantages of Ag-RDTs include low price, the lack of need for high-tech laboratory referral, and a short turnaround time to provide a result [2,3]. Although some contradictory findings have been reported, Ag-RDTs used as self-tests by the general population have a similar accuracy to when they are performed by health professionals [4–7].

Healthcare professionals, who are in contact with many patients, are at a high risk of exposure and, eventually if infected, of transmitting it to vulnerable patients. Above all, the most vulnerable ones such as oncological and immunosuppressed patients. The WHO recommends early detection of SARS-CoV-2 infection among health workers through syndromic surveillance and/or regular testing [7]. Little is known about the level of exposure of teachers and other professionals in the field of education, nevertheless the high transmissibility of the Omicron variant has also dramatically increased prevalence and incidence of SARS-CoV-2 in schools, where exposure is high as well.

The objective of the study was to implement and evaluate a pilot online offering of selftest kits for the SARS-CoV-2 rapid antigen screening test for healthcare and education professionals during the peak of the 6^th^ wave caused by the Omicron variant of SARS-CoV-2 in Catalonia (Spain).

## Methods

The study targeted two different key populations: 1) Staff of the Catalan Institute of Oncology (ICO), a public non-profit organization attached to the Catalan Health Service focused on cancer care and with 1,400 professionals distributed in 5 tertiary hospitals in Catalonia (Spain). 2) Staff of the schools belonging to the COVID Sentinel School Network of Catalonia (CSSNC), which monitors SARS-CoV-2 infection and its determinants, by means of repetitive cross-sectional surveys and includes 23 participating sentinel schools and 700 employees[8].

Data were collected prospectively during the peak of the 6^th^ wave due to the Omicron variant of SARS-CoV-2 in Catalonia, from 15 December 2021 to 15 February 2022.

Health care workers of the ICO and teachers from all participating schools of the CSSNC were invited to participate by email. Respondents accessed the study website (https://www.testate.org/), signed up and accepted through an online informed consent form. Then, participants requested a free COVID-19 rapid lateral flow home test kit (PanBIO ™ COVID-19 Antigen Self-Test, Abbot Laboratories, Chicago, US) and provided contact details including a postal address. Kits included a pictorial leaflet with guidance on how to perform the test and an instructional video was available on YouTube. After performing the test, participants completed an online survey including sociodemographic characteristics, clinical data, satisfaction with the intervention (Likert-type scale), ease of use of the kit, perceived advantages and disadvantages, result obtained, and a picture of the result. These pictures were assessed blind by the field coordinator and a microbiologist to check if the reading had been done correctly.

All participants with a positive result were contacted and were recommended both to self-isolate and to contact their General Practitioner (GP) as soon as possible.

We assessed the feasibility of the TESTATE COVID screening strategy among its users based on a conceptual framework adapted from earlier models [9,10]. The adapted framework divides the concept of feasibility into learnability, willingness, suitability, satisfaction, and efficacy.

The study was approved by the Ethical Committee of the Germans Trias i Pujol Hospital (PI-20-368).

## Results

During the study period 297 educators and 195 health professionals ordered a self-sampling kit, and 192 teachers (64.6%) and 111 health professionals (56.9%) correctly uploaded a picture of their result and answered the online survey (p: 0.1035). The median age of participants was 43.0 (IQR: 20.0-78.0), 80.1% were women.

We detected 18 positive cases (5.9%), including two cases which were identified as negative by the participants and as positive by the research team. The proportion of positive results was higher among teachers (7.3%) than health care professionals (3.6%) (p: 0.1959). Among positive participants there were: 11 teachers, three physicians, one nurse, two school administrative staff and one person without information. Most participants (78.5%) with a positive result had symptoms compatible with COVID-19 and all of them but two, contacted their GP and isolated themselves after knowing the result. One was not possible to contact and the other one ignored the recommendations because she considered herself to be negative, although the research team read the result as positive and informed her of it.

Most participants had received the COVID-19 vaccine (98.7%), among those vaccinated, 63.2% had had three doses and 30.0% had had two.

### Learnability

The majority of the participants (92.5%) found that the self-test was easy or very easy to use, 99.7% successfully completed the test, 88.9% did not need any help to perform the test and 93.5% agreed or strongly agreed with the statement “I trust that my interpretation of the result I obtained with the self-test is correct”, 81.6% of educators strongly agreed compared to 66.7% health professionals (P: 0.006) (Table 1). Two (0,6%) participants failed to read the test results.

**Table 1.**
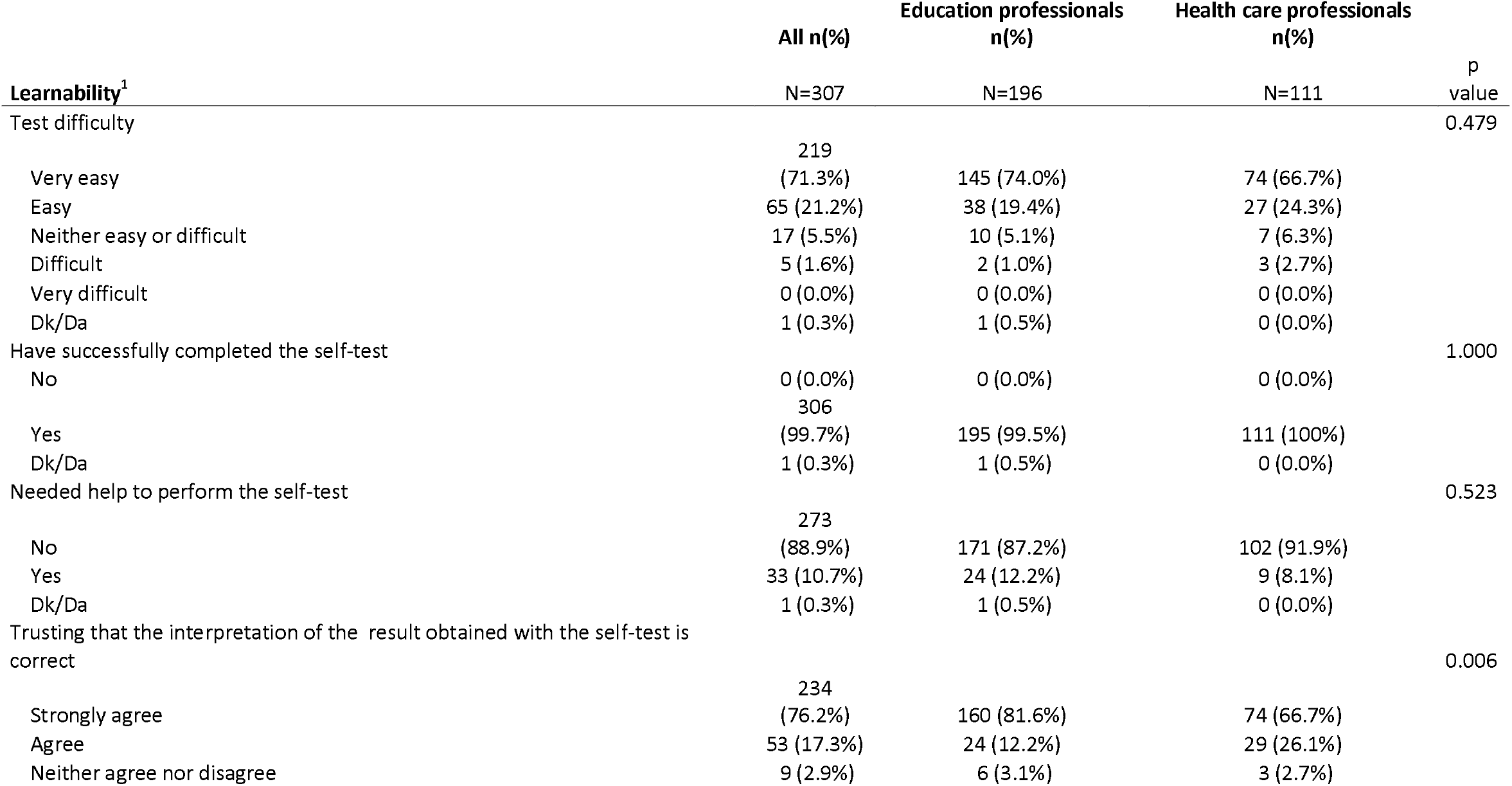
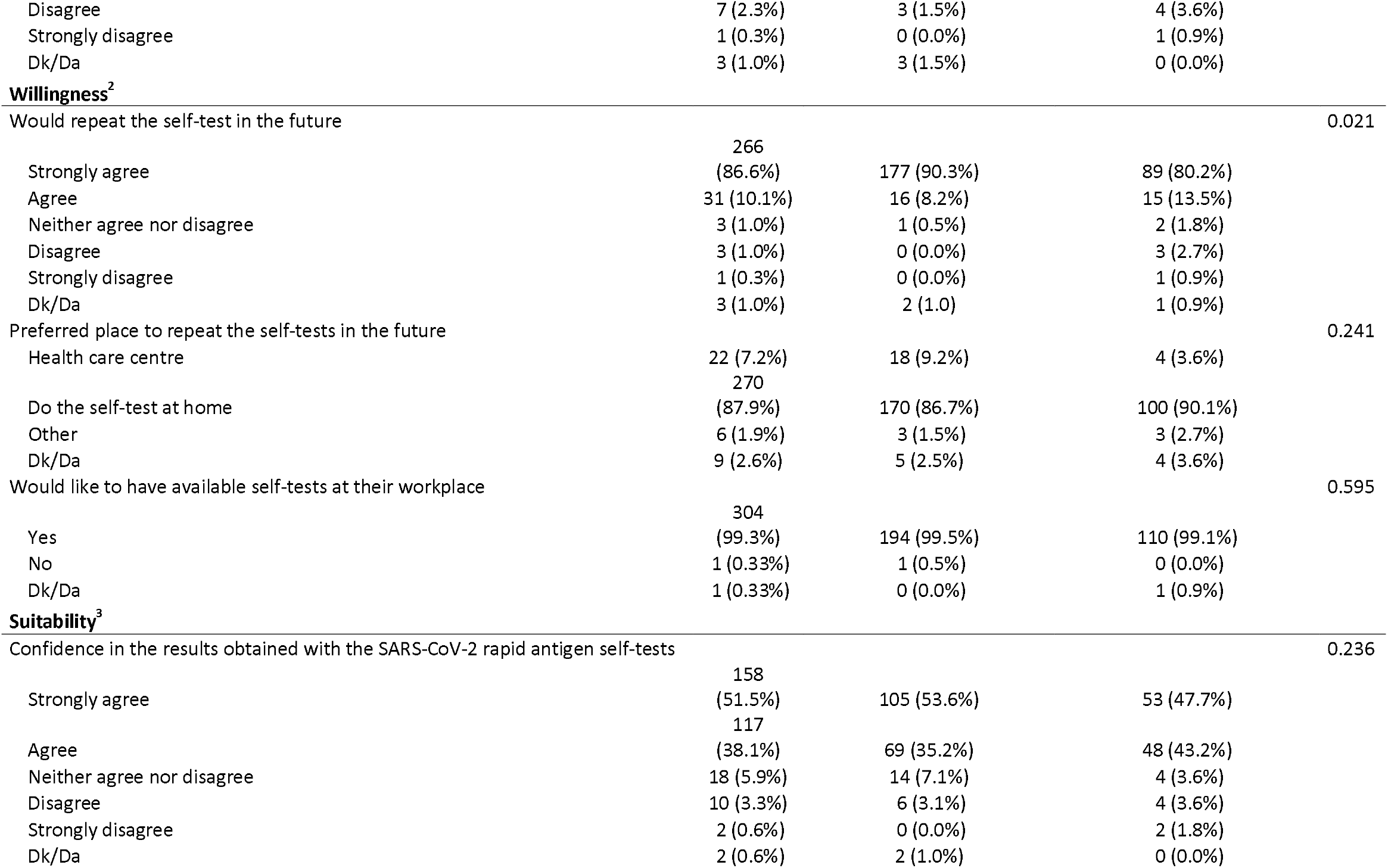
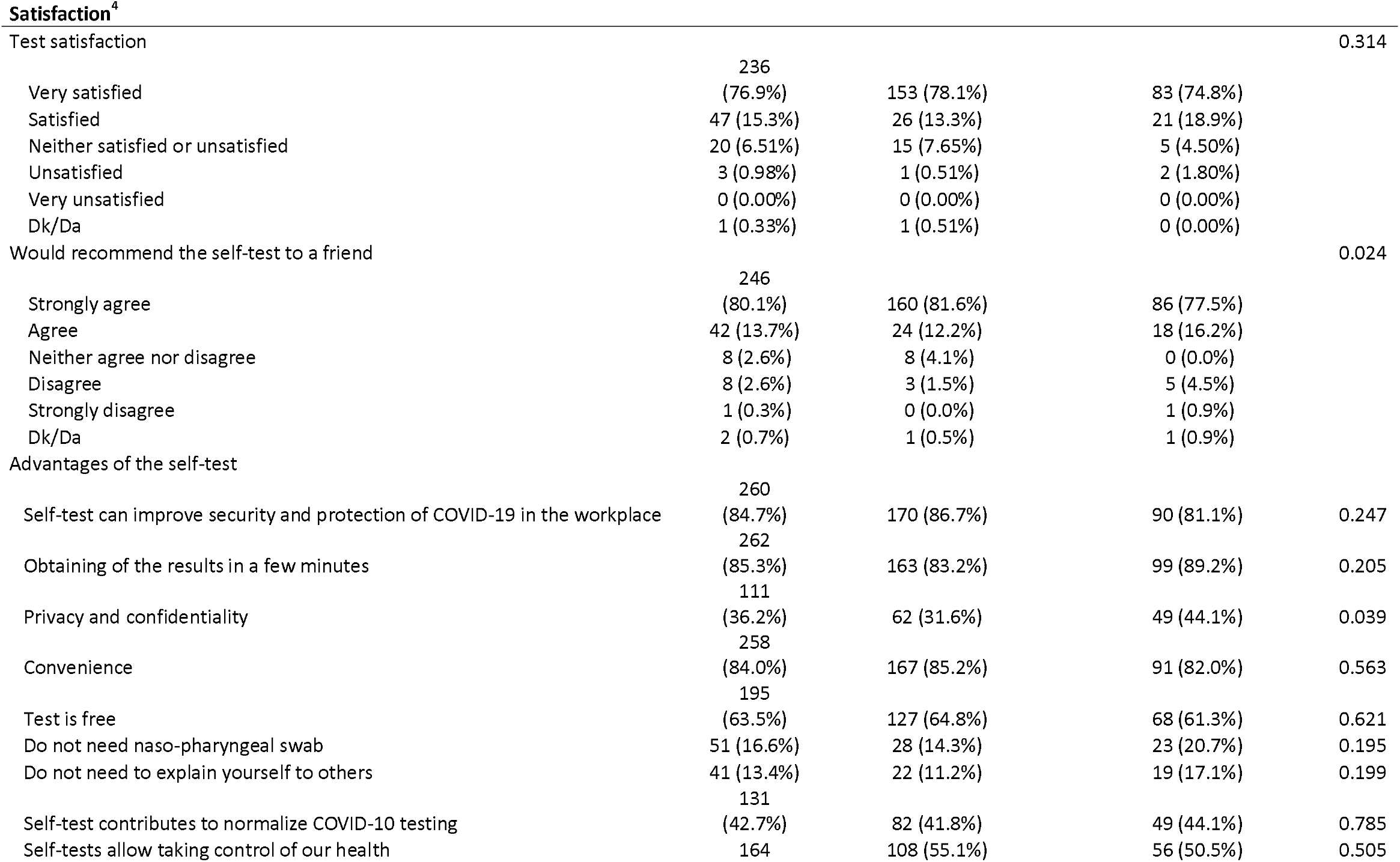
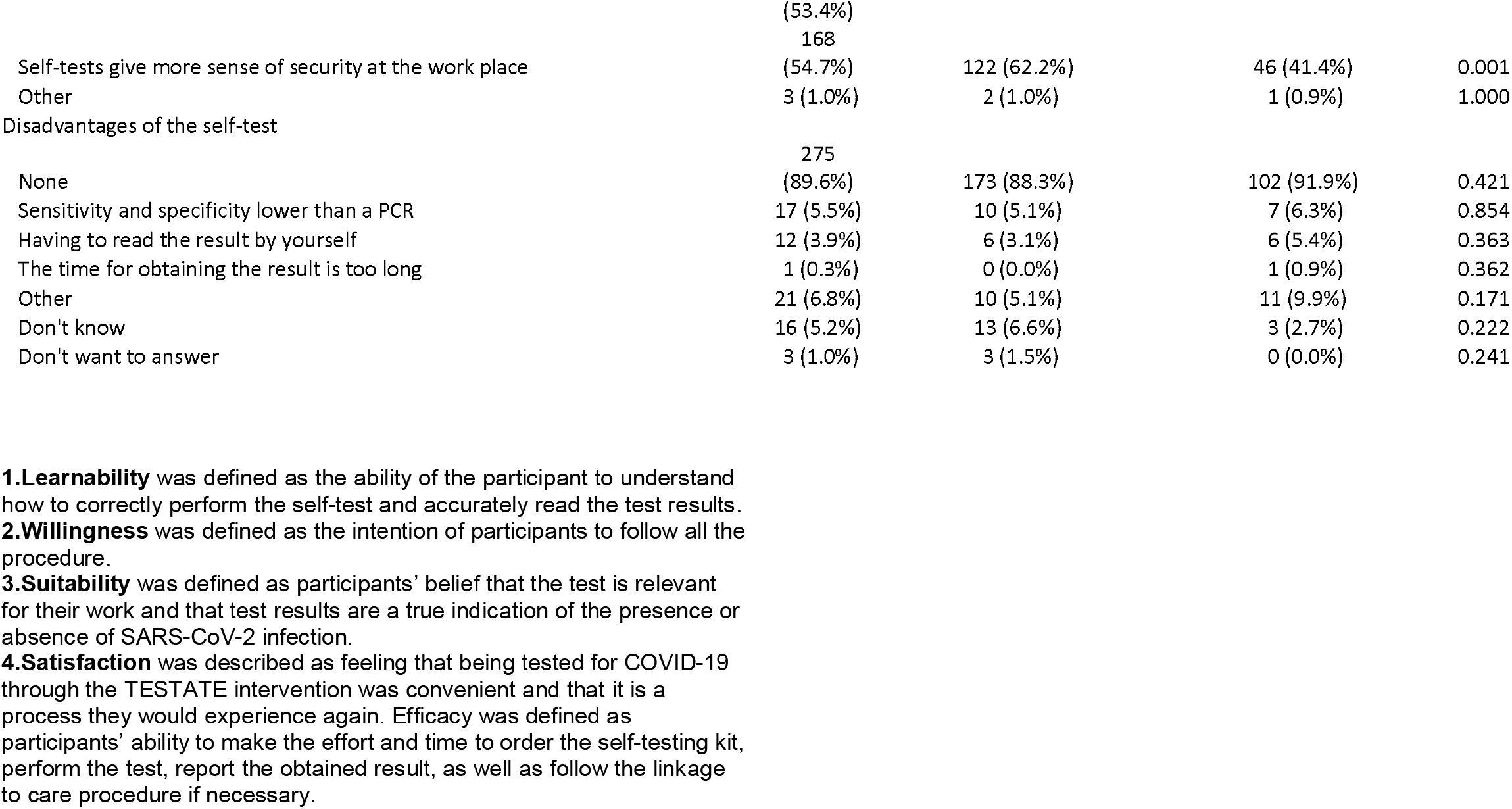
Learnability, willingness, suitability and satisfaction of the TESTATE COVID intervention addressed to health care and education professionals in Catalonia (Spain), N: 307. December 2021-February 2022.

### Willingness

Most of participants (96.7%) agreed or strongly agreed with the statement “I would repeat the rapid SARS-CoV-2 antigen self-test in the future”, 90.3% of educators strongly agreed compared with 80.2% of health professionals (p: 0.021). The most preferred way to repeat the test was “do the self-test at home” (87.9%); and 99.3% would like the test to be available at their workplace (Table 1).

### Suitability

Most participants (89.6%) agreed or strongly agreed with the statement “I trust the result obtained with the self-test” (Table 1).

### Satisfaction

92.2% of the participants answered that they were satisfied or very satisfied with the intervention; and 93.8% agreed or strongly agreed with the statement “I would recommend it to a friend” with 81.6% of educators strongly agreeing compared to 77.5% of health professionals (p :0.024). The advantages that were most identified were getting the result in a few minutes (85.3%) and the fact that the tests might improve safety and protection against COVID-19 at their workplace (84.7%); educators were more likely to identify “Self-tests give more sense of security at the workplace” as an advantage than health professionals (62.2% vs. 41.4%, p: 0.001); 89.6% did not identify any disadvantages.

## Discussion

Testing is a critical component of the overall prevention and control strategy for the COVID-19 pandemic[11]. Nevertheless, apart from contact tracing strategies, screening of key populations implies many logistical and operational challenges, including the necessity of periodic testing in periods of high incidence (ex. twice a week) in order to be effective [2]. Highly sensitive self-tests are cheap, simple, rapid tests and that can enable high frequency regimens that will capture most infections while they are still infectious.

Uploading the pictures of the results online contributed to better traceability of positives and this could improve the ability to break the epidemiological chain.

Although the study used an opportunistic sample, it shows the feasibility of implementation and provides an in-depth account of acceptability and usability of an online screening strategy based on antigen self-tests for COVID-19 addressed to health care and education professionals in Catalonia. This is the first time this was done in Spain, and during the peak of the 6^th^ wave caused by the Omicron variant. This information will be crucial to better define tailored screening strategies addressed to specific key populations, particularly during peaks of high community transmission of SARS-CoV-2 and eventually other respiratory transmitted agents.

## Data Availability

All data produced in the present study are available upon reasonable request to the authors

## Acknowledgments

The authors acknowledge the collaboration of Andreu Colom, Gema Ballega, Juan Rus, Marina Herrero, Pili Bonamusa, the General Direction of the Health Department and the Education Department of the Catalan Government of Catalonia, Anna Clopés, Deputy Director of the Catalan Institute of Oncology (ICO), Ana Sedano, Director of Human Resources of ICO and Abbot Laboratories.

